# Gene Portals: A Framework for Integrating Clinical, Functional, and Structural Evidence into Rare Disease Variant Classification

**DOI:** 10.64898/2026.03.05.26347086

**Authors:** Tobias Brünger, Ilona Krey, Suyeon Kim, Chiara Klöckner, Scott J Myers, Katrine M. Johannesen, Arthur Stefanski, Gary Taylor, Eduardo Perez-Palma, Marie Macnee, Stephanie Schorge, Rebekka S Dahl, Hongjie Yuan, Riley E. Perszyk, Sukhan Kim, Sunanjay Bajaj, Ingo Helbig, Jen Q. Pan, Mark Farrant, Lonnie Wollmuth, David J. A. Wyllie, Erkin Kurganov, David Baez, Sameer Zuberi, Christian M Boßelmann, Holger Lerche, Massimo Mantegazza, Sandrine Cestèle, Patrick May, Alina Ivaniuk, Mary Anne Meskis, Veronica Hood, Leah Schust, Kimberly Goodspeed, Jing-Qiong Kang, Amber Freed, Cornelius Gati, Ludovica Montanucci, Arthur Wuster, Marena Trinidad, Steven Froelich, Alexander T. Deng, Ángel Aledo-Serrano, Artem Borovikov, Artem Sharkov, Arjan Bouman, MJ Hajianpour, Deb K Pal, Leslie Danvoye, Damien Lederer, Tugce R. Balci, Eveline E O Hagebeuk, Alexis Heidlebaugh, Kathryn Oetjens, Trevor L Hoffman, Pasquale Striano, Sarah Drewes Williams, Kalene van Engelen, Katherine B. Howell, Jean Khoury, Tim A Benke, Vincent Strehlow, Konrad Platzer, Amy Ramsey, Lisa Manaster, Sunitha Malepati, Pangkong Fox, Jeffrey Noebels, Wendy Chung, Annapurna Poduri, Laina Lusk Stripe, Sarah M. Ruggiero, Stacey Cohen, Lacey Smith, Sylvia Boesch, Olivia Wilmarth, Anna Jenne Prentice, Esther Cha, Nikita Budnik, Marina P. Hommersom, Audra Kramer, Carlos G. Vanoye, Guo-Qiang Zhang, Michael Nothnagel, Aarno Palotie, Mark J. Daly, Alfred L. George, Yuri A. Zarate, Andreas Brunklaus, Stephen F Traynelis, Rikke S Møller, Johannes R. Lemke, Dennis Lal

## Abstract

Rare Mendelian disorders affect 300-400 million people globally. Although genetic testing has become widely adopted, gene-specific evidence for tailored variant interpretation remains scattered across resources. We present Gene Portals, a framework for gene-centered multimodal knowledge bases that co-localize expert-harmonized clinical data, functional assays, population variation, structural annotations and gene-specific ACMG/AMP specifications within a single resource. A modular interface integrates this unified evidence with VCEP-refined ACMG specifications to enable automated gene-specific variant classification, infer molecular mechanisms, and support cross-gene analyses. We demonstrate the framework’s utility across five Gene portals spanning eleven neurodevelopmental disorder-associated genes, integrating data from 4,423 individuals with 2,838 unique variants, 36,149 ClinVar submissions, and 1,044 expert-curated molecular readouts. By organizing evidence that is otherwise dispersed across multiple sources into a unified, queryable framework, the SCN, GRIN, CACNA1A, SATB2 and SLC6A1 Gene Portals became widely used community resources and provide an extensible template for standardized rare-disease variant interpretation and mechanism-aware discovery.

## Main

More than 7000 rare Mendelian disorders affect 300-400 million individuals gloabally. However, even when the causal gene is known, allelic heterogeneity and pleiotropy generate substantial clinical and mechanistic diversity: different pathogenic variants in the same gene can alter dosage, biophysical properties, protein stability, trafficking, protein-protein interactions, or cell-type–specific function in distinct ways. In genes encoding ion channels, receptors, and transcriptional regulators, these diverse molecular effects, such as loss, gain or mixed function, are associated with broad phenotypic spectra that often blur classical syndromic labels^1–3^. As a result, accurate variant classification increasingly requires integrating clinical evidence with variant-specific functional and protein-specific structural data, yet the tailored data remain fragmented and difficult to access for affected individuals, clinicians, genetic counselors, and researchers.

Current resources for variant classification are characterized by two related limitations. First, clinically well-characterized patient cohorts and targeted functional assay data are typically generated, curated, and disseminated in gene- or study-specific efforts, often using heterogeneous formats and vocabularies, with limited harmonization across datasets. Second, even when high-quality data are available, clinical, population, functional, structural, and *in silico* evidence are typically organized in isolation rather than interconnected within a unified gene-centered framework. As a result, existing platforms capture only isolated components of the broader evidence landscape and rarely support systematic integration of phenotypes linked to individual variants, mechanistic functional evidence, structural context, and gene family-aware information^4–8^. Genome-wide tools^9,10^ implementing the American College of Medical Genetics and Genomics and American Molecular Pathology (ACMG/AMP) ^11^ guidelines provide standardized and scalable support for variant classification across large datasets. However, their reliance on generic, large-scale evidence resources and limited support for gene-tailored datasets and annotations may be insufficient for rare-disease genes in which variant pathogenicity and clinical presentation are tightly coupled to positional context and molecular disease mechanisms^12^.

Together, these constraints highlight the need for a gene-centric framework that integrates clinical, functional, structural, and population-level evidence within a unified resource to enable tailored data interpretation at single-variant resolution. To address this gap, we developed the Gene Portal (GP) framework, which aggregates and harmonizes multimodal datasets, links variants across transcripts and gene families, and contextualizes evidence along protein sequence and three-dimensional structure. The framework supports both expert-guided and automated ACMG classification and is designed to incorporate continuously updated clinical and experimental data within a centralized infrastructure.

As proof of concept of the framework, we implemented it across five GPs encompassing eleven neurodevelopmental disorder (NDD) associated genes. By transforming previously fragmented datasets into interoperable, mechanism-aware knowledge bases, this implementation establishes a scalable architecture for standardized variant interpretation and cross-gene genotype–phenotype analyses.

## Results

### A multimodal platform integrating clinical, genetic, structural, and functional evidence

Each GP is built on a gene-centered knowledge base that provides synchronized access to aggregated and harmonized multidomain datasets at single-variant resolution (Fig. 1). For each gene, this knowledge base includes expert-curated clinical phenotypes and functional assay results from peer-reviewed literature. Collaborator-contributed datasets are also integrated and, when unpublished, undergo independent review by multiple external domain experts to confirm data quality and interpretation before inclusion. For a given gene or gene family, variant-level data are represented as unified entities mapped across transcripts, protein isoforms, paralogous genes, and three-dimensional structures, enabling evidence from multiple domains to be interpreted in a shared positional and mechanistic context. Users can interact with the knowledge base through three main modules. The *Clinical Overview* module allows users to examine aggregated cohort characteristics, including gene-specific phenotype spectra and comorbidities. The *Variant C*lassification module enables users to evaluate individual variants using gene- and variant-tailored ACMG/AMP¹² classification while assessing experimentally derived and/or predicted effects on protein function. The *Research* module allows users to explore linked clinical and functional evidence across genes and to visualize variant distributions along linear protein sequences and within three-dimensional protein assemblies.

**Figure 1.**
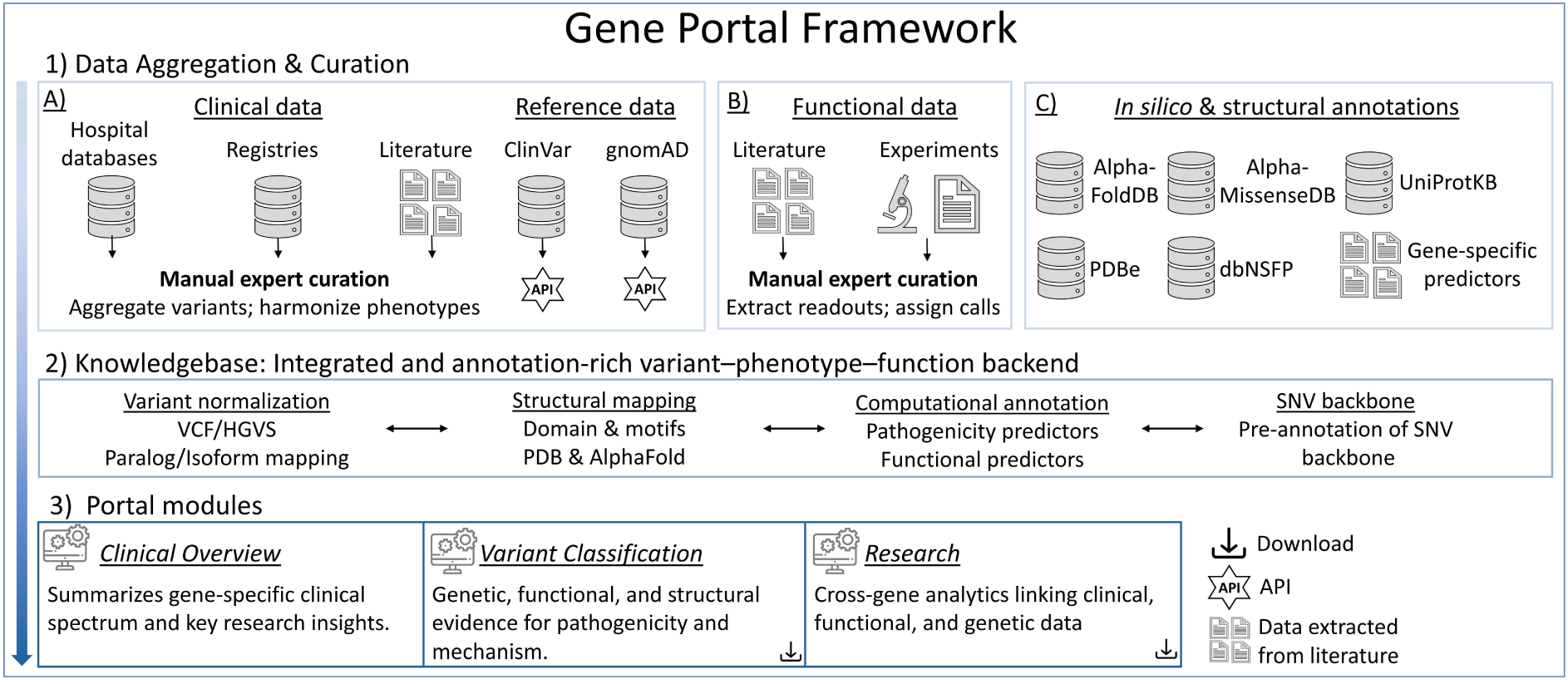
Architecture of the Gene Portals (GP) framework integrating clinical, genetic, functional, and structural evidence. The GP unifies curated clinical, population, functional, and *in silico* data into a standardized, gene-specific knowledge base. **Panel 1** summarizes data aggregation across three evidence domains: (A) clinical and population data comprising expert-curated patient cohorts, literature-derived cases, registry datasets, and variant submissions from ClinVar and gnomAD; (B) functional data capturing electrophysiological and other molecular readouts extracted from published and unpublished experiments (see Methods for review details); and (C) computational and structural annotations integrating protein features, AlphaFold and PDB structures, genome-wide prediction scores, and gene-specific functional predictors. **Panel 2** depicts the unified knowledge base backend that links variant, phenotype, functional, and structural information at single-variant resolution. The backend is generated through a standardization and annotation workflow performing VCF and HGVS normalization, isoform and paralog mapping, domain and motif alignment, three-dimensional structure integration, application of computational evidence, and construction of a comprehensive annotated coding SNV backbone. **Panel 3** presents the interactive GP modules: The *Clinical Overview* module summarizes gene-specific phenotypes, comorbidities, and cohort characteristics. The *Variant Classification* module integrates clinical, functional, structural, and population evidence to support automated ACMG classification using gene-tailored criteria. The Research module enables cross-gene exploration of variant–phenotype–function relationships, residue-level mutational patterns, and structural context. Together, these components provide a reproducible and extensible system for mechanism-aware variant classification across rare-disease genes. **Abbreviations:** ACMG: American College of Medical Genetics and Genomics; API: Application programming interface; PDB: Protein Data Bank; HGVS: Human Genome Variation Society nomenclature; VCF: Variant Call Format; SNV: Single Nucleotide Variant.

This framework was first applied to N-methyl-D-aspartate receptor (NMDAR) encoding genes, creating the *GRIN* portal. At present, there are five independently deployed GPs across eleven clinically and mechanistically heterogeneous NDD-associated genes: the *SCN* (encoding voltage-gated sodium channels), GRIN (encoding N-methyl-D-aspartate receptors, NMDAR), *CACNA1A* (encoding the CaV2.1 calcium channel), *SATB2* (encoding DNA-binding protein SATB2), *and SLC6A1* (encoding the GABA transporter protein type 1) GPs, which are all publicly accessible at https://lalresearchgroup.org. User surveys show that clinicians and researchers report accelerated variant review and additional decision-relevant context, and web analytics demonstrate rapid community uptake with an average of >600 monthly active users globally over a six-month period (Supplementary Data ‘User Feedback and Global Reach’, Supplementary Figure 1). Regular, versioned updates integrate new clinical reports and functional studies to expand coverage over time.

### Resources in the Gene Portals

#### Integrated data landscape

In their current deployment, the GPs integrate expert-curated clinical information from 4,423 affected individuals, 2,838 unique variants, 36,149 ClinVar submissions, and functional readouts for 1,044 variants across 11 genes. Although ClinVar represents the largest single publicly available collection of patient variants for these genes, only 6,574 variants (18.2%) are classified as likely pathogenic or pathogenic across all genes, whereas 24–58% of ClinVar submissions are variants of uncertain significance (VUS) (Fig. 2B). Against this background, data from clinical research networks substantially expand the spectrum of represented variants; for example, 36.2% (N = 603) of missense variants observed in the curated patient cohorts are absent from ClinVar’s list of likely pathogenic and pathogenic variants. The list of expert-curated variants reveals marked gene-specific profiles: missense variants dominate in *SLC6A1*, most voltage-gated sodium channel genes (*SCN* genes), and most *N*-methyl-d-aspartate-type glutamate receptor genes (*GRIN* genes), whereas null variants are more frequent among individuals with *SATB2* variants (Fig. 2A). A central principle of the GP framework is that variant evidence is contextualized at the level of conserved protein positions, which is already implemented in the ACMG/AMP variant curation criteria in epilepsy-related sodium channels^13^. For the GP framework, we have applied this concept across gene families of related (paralogous) genes more broadly: (Likely)-pathogenic variants at a given amino acid residue can provide evidence for the variant classification of other substitutions at the same position, and homologous residues in paralogous genes provide evidence when sequence and structural constraints are conserved^1,11,12^. Incorporating mappings on residue-level and across paralogs increases the fraction of missense SNVs that can be informed through established patient variants from 2% to as much as ∼30% (Fig. 2C). Complementary, functional assays are available for only 0.2–3.8% of possible missense SNVs, yet positional and paralog-based mapping extends the fraction of all missense SNVs that available functional assays can support up to 23% (Fig. 2D). In addition, to capture broader sequence patterns from curated patient variants, we computed pathogenic variant–enriched regions (PERs)^14^, which span 1.5–18.5% of residues across genes (Fig. 2E) and highlight domains enriched for curated and ClinVar likely pathogenic and pathogenic missense variants relative to gnomAD.

**Figure 2.**
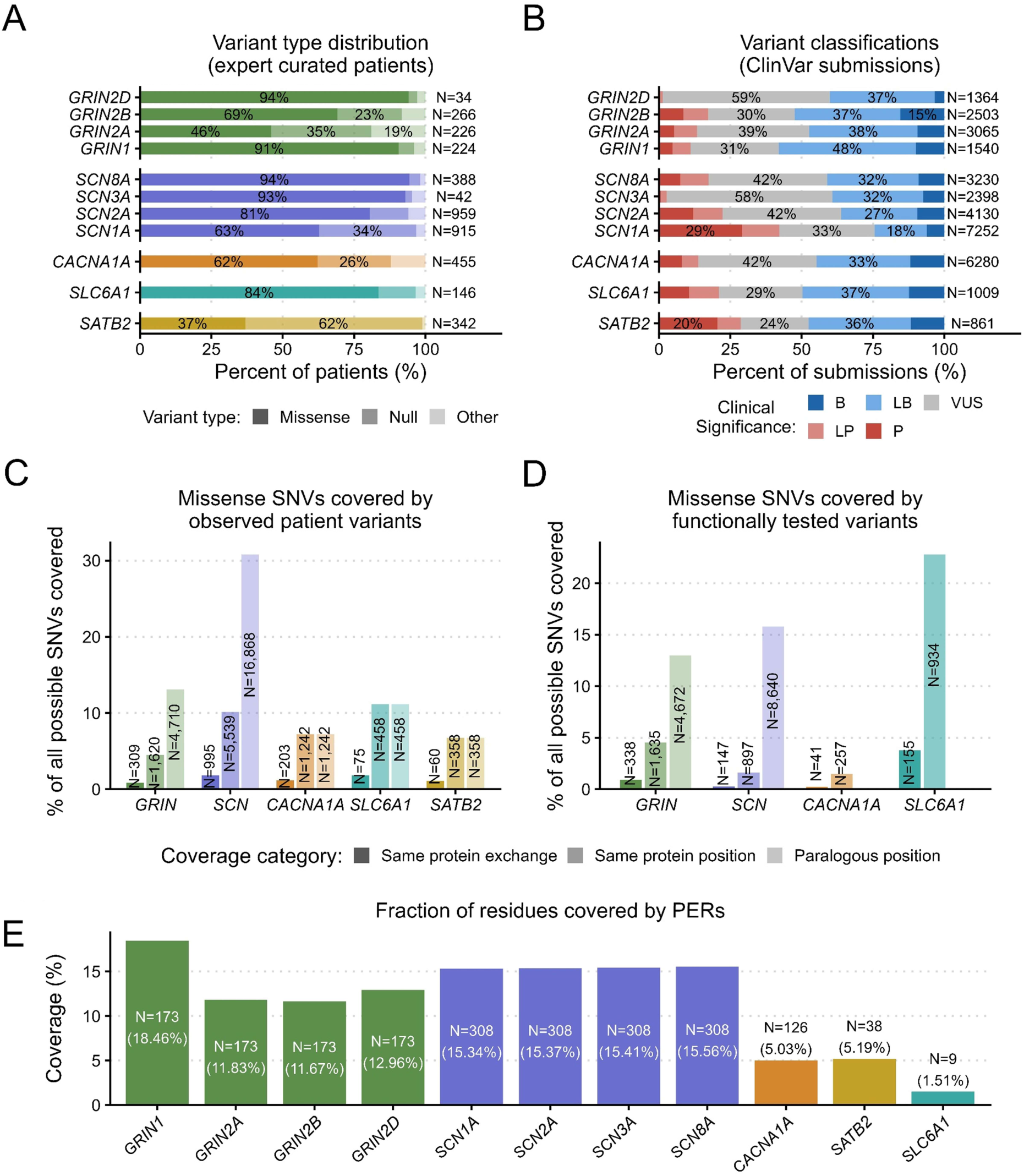
Composition and coverage of integrated clinical, ClinVar, and functional datasets. **(A)** Variant-type distributions are shown for expert-curated individuals across 11 genes.**(B)** Clinical significance of 36,149 ClinVar submissions harmonized in the shared schema, showing 35–60% uncertain/conflicting classifications across genes. **(C)** Fraction of all possible missense SNVs located at positions represented by curated patient variants, shown as coverage of the same residue, same amino-acid exchange, or paralog-aligned position. **(D)** Equivalent coverage based on functionally tested variants highlights the smaller but complementary sequence space informed by experimental assays. **(E)** Considering the MANE transcript-associated isoform, we calculated the fraction of residues located in pathogenic variant–enriched regions (PERs). PERs cover 1.5–18.5% of coding residues across genes. N indicates the number of residues per gene that fall within PERs. **Abbreviations**: P: Pathogenic; LP: Likely-pathogenic; VUS: Variant of uncertain significance; LB: Likely-benign; B: Benign; Null: nonsense, frameshift, canonical splice-site, and start-loss variants; Other: non-missense and non-null variants, including in-frame indels and multigene copy-number variants.

#### Integrated pathogenicity and functional evidence

The harmonized GP backend also enables systematic evaluation of pathogenicity metrics and gene-level functional prediction models across gene families. The distributions of six widely used variant pathogenicity classification scores (AlphaMissense^15^, REVEL^16^, CADD^17^, EVE, PARA-Z^18^, and MTR^19^) differ between *CACNA1A*, disease-associated *GRIN* genes (*GRIN1*, *GRIN2A*, *GRIN2B*, *GRIN2D*), *SCN* genes (*SCN1A*, *SCN2A*, *SCN3A*, *SCN8A*), *SATB2*, and *SLC6A1*, and across expert-curated patient, ClinVar, and gnomAD variants (Fig. 3). These patterns highlight strong gene- and score-specific variation in the ability to distinguish presumably pathogenic and benign variants. In each GP, the distributions of prediction scores for patient-derived and population reference variants within the same gene are displayed alongside the user-selected variant, providing visual context to guide the selection and interpretation of computational evidence for variant classification.

**Figure 3.**
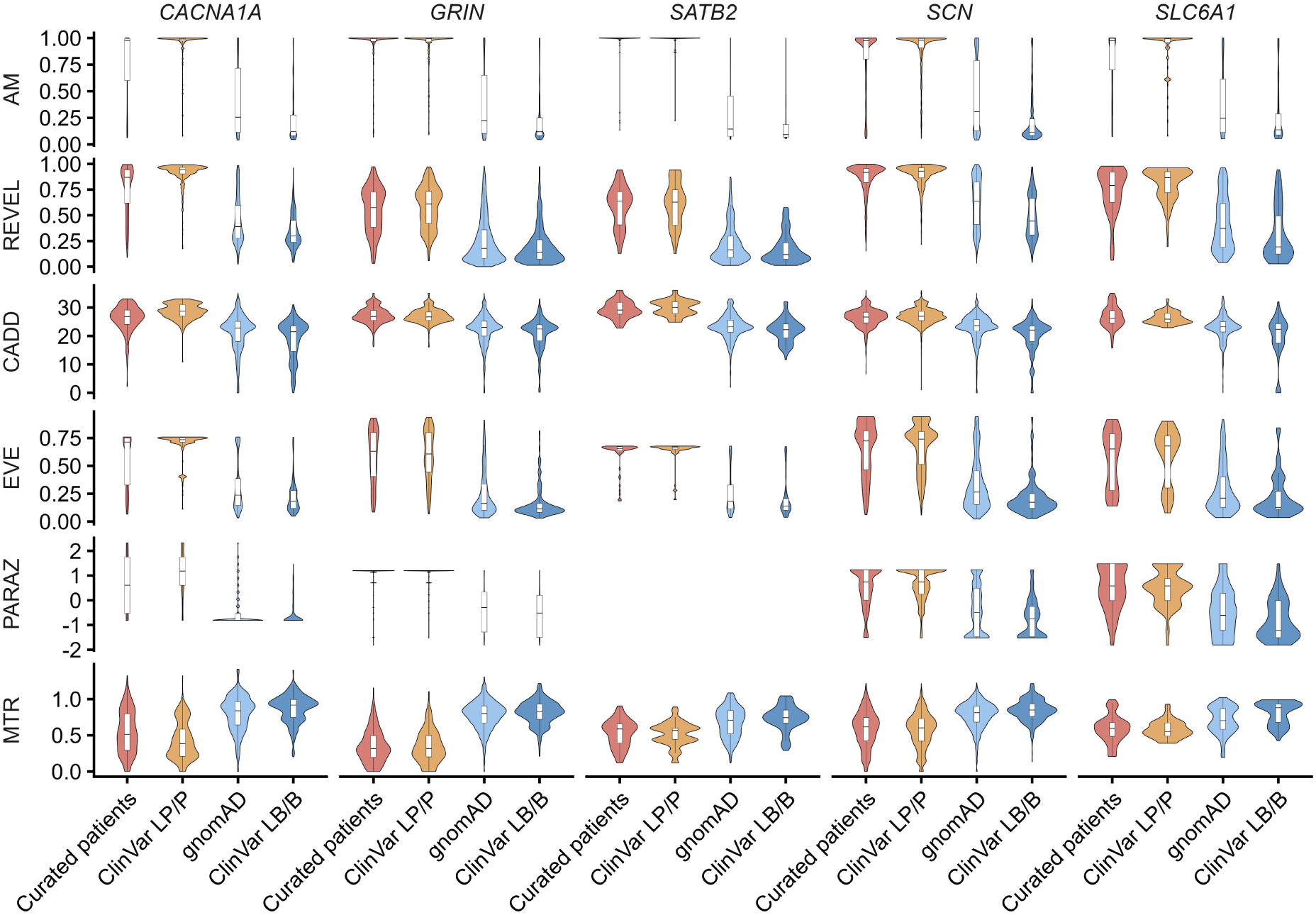
Pathogenicity score distributions across Gene Portals (GPs). Distributions of six widely used pathogenicity and constraint scores (AlphaMissense, REVEL, CADD, EVE, PARA-Z and MTR) across curated patient variants, ClinVar pathogenic/likely pathogenic and benign/likely benign variants, and control variants from gnomAD for the four gene families represented in the GPs. The patterns reveal gene- and score-specific separability between presumably pathogenic and benign variants. **Abbreviations**: P: Pathogenic; LP: Likely-pathogenic; LB: Likely-benign; B: Benign.

### Gene Portal modules

All GPs implement a common interface architecture design with three core analytic modules - *Clinical Overview* (CO), *Variant Classification* (VC), and *Research* (R), ensuring consistency and comparability across genes while allowing gene-specific customization. In addition, selected GPs include two optional modules focused on education and prospective data collection, the *Educational Resources* module and the *Registry* module. Layout, visualizations, and available filters are adapted to the structure and completeness of the underlying datasets and refined in collaboration with scientific, clinical, and family communities. This combination of a standardized backbone layer with disease community-informed customization ensures methodological coherence across GPs while allowing each GP to remain responsive to the needs of its respective disease community. The GPs are already endorsed by 55 patient advocacy groups (PAGs) (Supplementary Table 1), underscoring broad support for their flexible application.

#### Module I – Clinical Overview (CO)

The CO module illustrates how gene-specific clinical knowledge is synthesized into an interpretable summary for both experts and non-experts. For example, in the *CACNA1A* portal, a timeline aggregates key milestones in *CACNA1A* research from the earliest gene–disease association studies implicating *CACNA1A* in familial hemiplegic migraine and episodic ataxia to more recent genotype–phenotype and natural-history studies that delineate clinical subtypes (Fig. 4A). The module further provides a structured overview of *CACNA1A*-related phenotypes (Fig. 4B). All reported features from the curated cohort and literature are standardized to Human Phenotype Ontology (HPO) terms^20^, enabling quantitative comparisons across variant classes. In the registry-based clinical summary (Fig. 4C), missense variant carriers are enriched for epilepsy and global developmental delay, whereas protein-truncating variants are more frequently associated with ataxia and nystagmus. Together, these views demonstrate how the CO module converts heterogeneous registry and literature data into a gene-specific, variant-type–aware disease profile that can be rapidly inspected in clinical practice.

**Figure 4.**
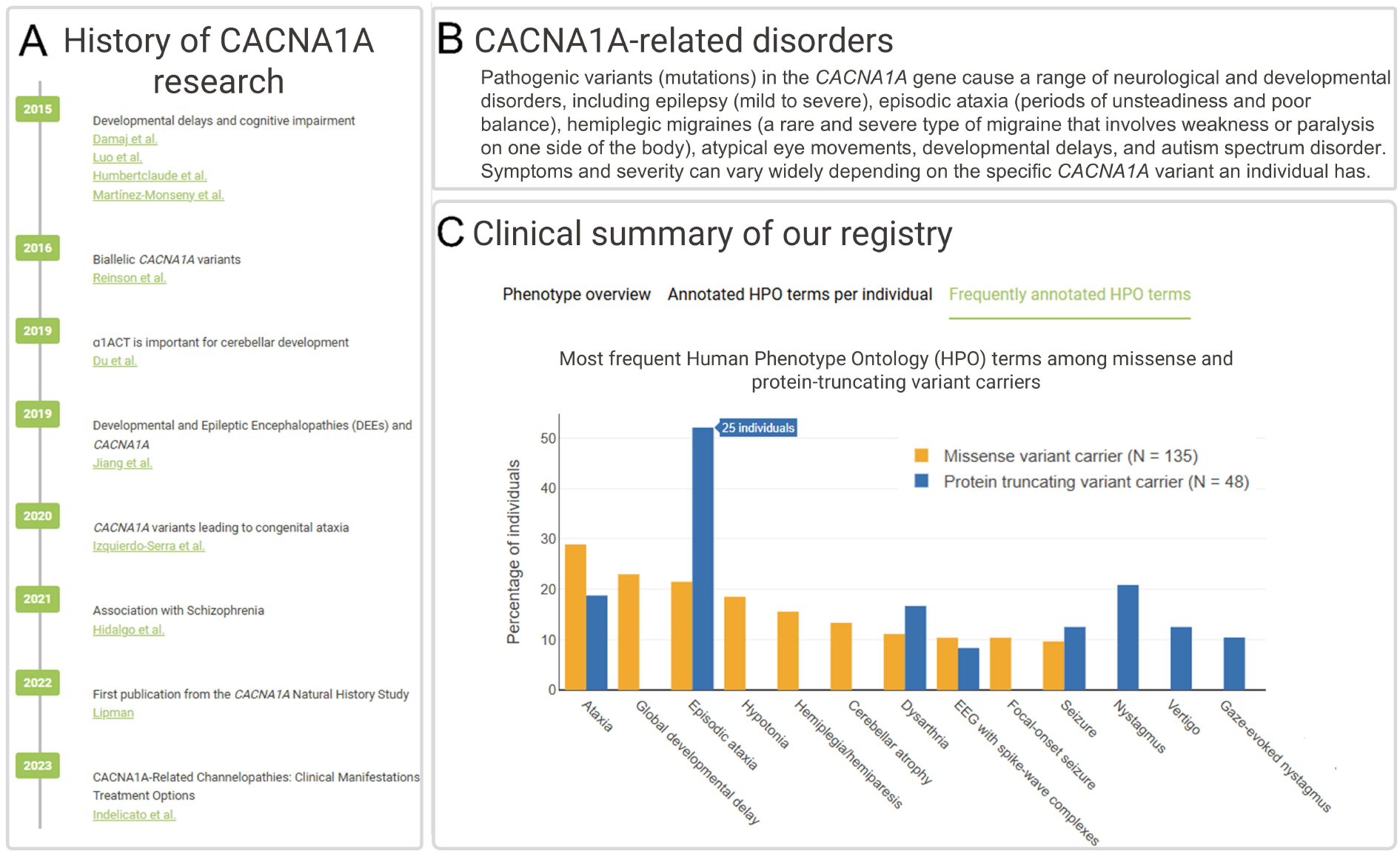
Structure and key features of the Clinical Overview module illustrated for the *CACNA1A* portal. **(A)** Timeline summarizing major milestones in *CACNA1A* research, from early gene-disease association studies to recent genotype-phenotype analyses refining clinical subtypes. **(B)** Overview of *CACNA1A*-related disorders outlining the major neurological and developmental phenotypes, including episodic ataxia, developmental and epileptic encephalopathy, hemiplegic migraine, and autism spectrum disorder. **(C)** Clinical summary displaying the most frequent Human Phenotype Ontology (HPO) terms observed among individuals with *CACNA1A* variants, illustrating differences in phenotype prevalence between missense and null variant carriers.

#### Module II – Variant Classification (VC)

To demonstrate the functionality of the VC module, we highlight a representative example from the *GRIN* portal (*GRIN2B (*NM_000834.5): p. Gly820Ala. Users start by specifying gene, transcript, and variant at the cDNA or protein level (Fig. 5A). The *GRIN portal* then returns a variant summary including transcript context (NM_000834.5) and an automated ACMG/AMP classification, which is pathogenic in this case (Fig. 5B). This classification is generated directly from co-localized, expert-harmonized clinical and functional datasets integrated within the *GRIN portal*, that are otherwise unavailable, eliminating the need to retrieve evidence from disparate external sources. Notably, several classification criteria have been refined for the GRIN genes according to the currently preliminary specifications of the ClinGen GRIN Variant Curation Expert Panel (VCEP)^21^, which will be released soon. Each applied criterion is explicitly listed with its assigned weight, and users can immediately export a structured HTML report for documentation or multidisciplinary review. The evidence supporting this classification can be explored across several coordinated panels: In the example shown in Figure 5, a cohort table summarizes six individuals from the Global GRI Registry carrying p.(Gly820Ala) and lists their associated phenotype summaries, while cross-gene sequence alignments identify additional patients with variants at paralogous conserved alignment index positions in *GRIN1*, supporting application of PS1 and related criteria^12^ (Fig. 5C). Functional data visualizations aggregate six experimentally measured molecular parameters that have been selected for clinical interpretation by the ClinGen GRIN VCEP^22^, indicating a likely loss-of-function (LoF) effect consistent with reduced open probability and altered kinetic properties (Fig. 5D). These results align with the predictions from a GRIN-specific variant function prediction model^23^, providing orthogonal support for the ACMG/AMP computational evidence criterion (PP3). *In silico* scores, including a high REVEL value, separate patient and population variants and support PP3, while the variant location within a pre-computed mutational hotspot displayed in both linear protein sequence and 3D receptor structure contributes to the mutational hotspot criterion (PM1) (Fig. 5E). In addition to mutational hotspots, the *GRIN portal* displays three-dimensional missense intolerance (3D-MTR), highlighting structurally constrained regions of the protein. By integrating these data into a single interactive view and mapping them directly onto ACMG/AMP criteria in accordance with the most recent VCEP specifications, the VC module allows users to move seamlessly from variant query to transparent, reproducible, gene-tailored classification. The utility of this module for ongoing research efforts is highlighted by a recently published prediction model to predict the functional effects of missense variants in *GRIN* genes^23^, which was built using data from *GRIN p*ortal. The GRIN functional predictions are also accessible through the *GRIN portal*.

**Figure 5.**
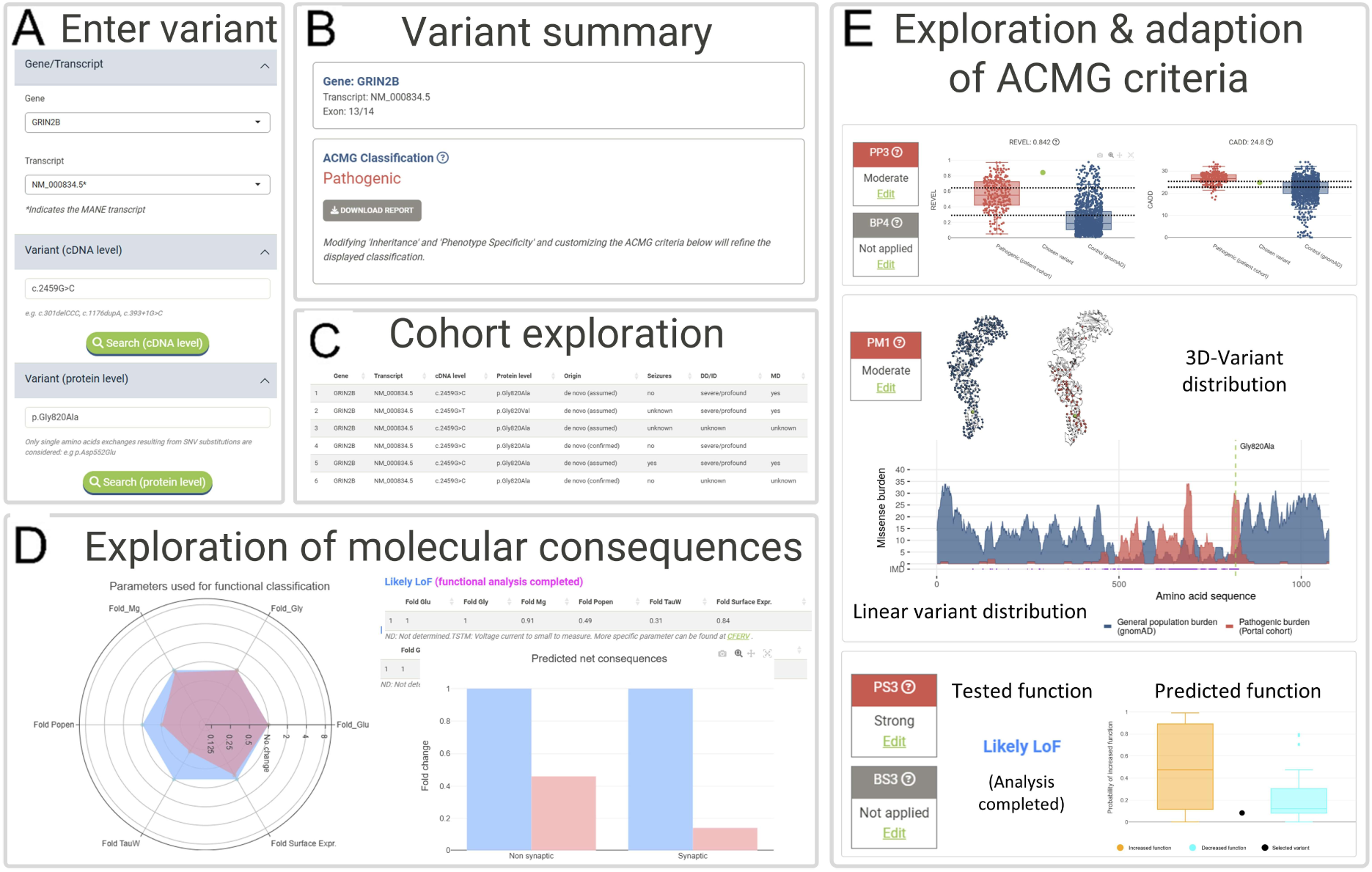
Structure and key functionalities of the Variant Classification module exemplified by the *GRIN portal*. **(A)** Variant input panel where users can search by gene, transcript, and variant at the cDNA or protein level. **(B)** Automated ACMG classification summary for the *GRIN2B* variant (NM_000834.5:p.Gly820Ala) with an option to export a detailed HTML report (see Supplementary data “GRIN Portal: Variant Analysis Report”). **(C)** Cohort exploration table showing six expert-curated individuals from the Global GRI Registry carrying the same variant and their associated clinical features. **(D)** Functional data visualization summarizing experimentally tested molecular parameters used for classification, indicating a likely loss-of-function (LoF) effect of the N-methyl-D-aspartate receptor (NMDAR). **(E)** Interactive ACMG criterion exploration with data-supported evidence, including pathogenicity predictor plots (PP3), mutational hotspot mapping (PM1), and integrated variant distributions on the linear protein sequence and protein structure alongside tested and predicted functional outcomes.

#### Module III – Research (R)

The R module demonstrates how the GPs support hypothesis generation using linear protein sequences, 3D structures, and biomedical data visualizations. In the *GRIN portal*, users can filter by gene, variant type, location within a mutational hotspot, functional consequence, and phenotype using a flexible query interface (Fig. 6A). For example, selecting variants annotated as “likely loss-of-function (LoF)” across the *GRIN1*, *GRIN2A*, and *GRIN2B* cohorts reveals distinct clustering patterns along the linear protein sequence, with most LoF variants localizing in mutational hotspots (Fig. 6B). The same variants are projected onto the NMDAR complex structure, enabling visual assessment of whether LoF variants preferentially affect specific domains, interfaces, or subunits. The phenotype interface links these molecular patterns to clinical outcomes (Fig. 6C). Individuals with missense LoF variants in *GRIN1* predominantly exhibit severe intellectual disability, whereas those with missense LoF variants in *GRIN2A* or *GRIN2B* span a broader range of cognitive severity. Seizure frequency also differs by gene: more than 80% of individuals with missense LoF variants in *GRIN2A* have seizures, compared with approximately 20% of those with missense LoF variants in *GRIN2B*. These cross-gene views illustrate how the R module connects variant location, known functional consequences, structural context, and phenotype, enabling domain-level hypotheses about mechanisms and their correlations with clinical phenotypes and function.

**Figure 6.**
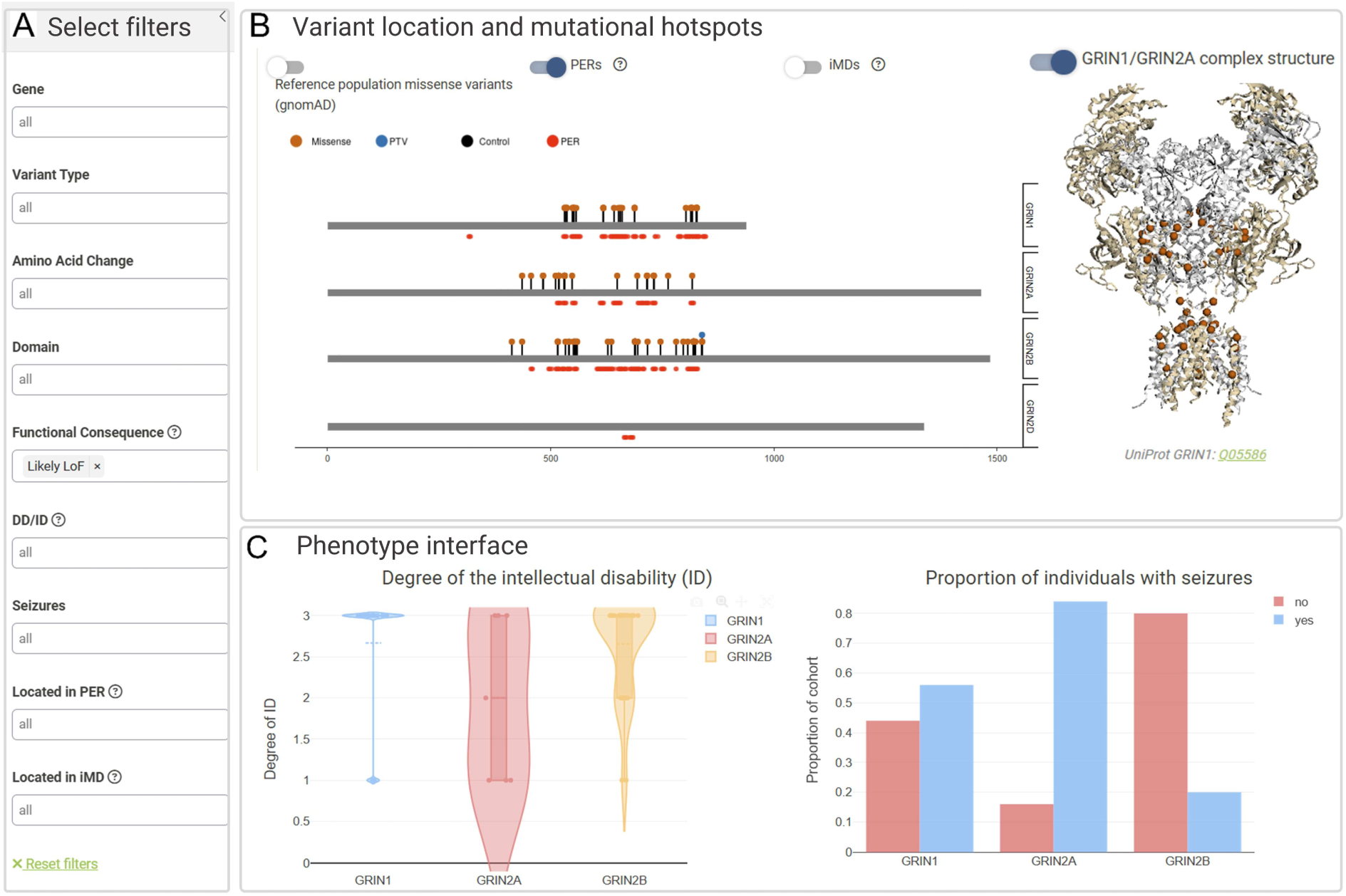
Structure and functionalities of the Research module illustrated for the *GRIN* portal. **(A)** Filtering panel allowing selection by gene, variant type, mutational hotspot, functional consequence, or phenotype. **(B)** Distribution of likely loss-of-function (LoF) variants mapped onto the linear protein sequence with pathogenic variant–enriched regions (PERs) and visualized on the GLuN1-GluN2A (*GRIN1*, *GRIN2A)* receptor complex structure (PDB ID: 6MMB). **(C)** Phenotype interface showing degree of intellectual disability and seizure frequency among individuals with LoF variants across *GRIN1*, *GRIN2A*, and *GRIN2B*.

#### Module IV – Educational Resources

Most GPs include an educational resource module designed to lower the barrier to entry for clinicians, researchers, trainees, affected individuals, and families. For families, these resources facilitate the return and contextualization of genetic findings by translating complex molecular and clinical concepts into accessible explanations, thereby supporting informed decision-making and sustained engagement. This module provides expert-designed educational videos and tutorials that explain the genetic architecture of each disorder, the principles of functional assays, and key aspects of variant classification. Subtitles in multiple languages enhance accessibility for international users and patient communities. GPs also include, if available, curated links to relevant patient advocacy groups, facilitating bidirectional communication between clinicians, researchers, and affected families.

#### Module V – Registry

Both the *GRIN* and *SATB2* GPs include an integrated registry module that provides families with easy access to, and enrollment and participation in research directly through the GPs. As an illustrative example, the *GRIN portal* implements a REDCap-based Global GRIN Registry that captures harmonized genetic and phenotypic information across a broad range of disease features. After enrollment, data will be curated and validated, and if questions or inconsistencies arise, families will be re-contacted to request additional information, if necessary. The registry currently includes data from 773 affected individuals, and participants may be re-contacted for eligibility screening in ongoing research studies, natural history efforts, or interventional trials. Thus, the GP is not only a repository for newly generated clinical and functional data to be aggregated, curated, and used in research projects, but also coordinates clinical trial readiness through registries maintained by patient advocacy organizations. The registry data of Module V feeds back the clinical overview displayed in Module I after an additional curation for quality assurance.

Particularly, modules IV and V extend the framework from a static knowledge base to a living ecosystem that supports education, community engagement, and prospective data collection.

## Discussion

Genomic resources such as ClinVar^5^, HGMD^4^, and DECIPHER^7^ catalog disease-associated variants, while others, such as MaveDB^8^, list variant consequences quantified in high-throughput assays. These resources provide indispensable references at population or genome-wide resolution. However, for most rare-disease genes, the underlying clinical and experimental evidence remains fragmented across case reports, patient registries, and isolated functional studies, which are inconsistently formatted and often difficult to access and integrate. Patient registries are a particularly valuable, yet underutilized, source of variants not uploaded to ClinVar by diagnostic laboratories (36.2% of missense variants in our resources), as well as rich longitudinal and deeply phenotyped clinical data. GPs address this gap through community-driven, gene-centered knowledge bases that interconnect expert-curated clinical phenotypes, functional assay data, and structural annotations, alongside population references, within unified, variant-centric, modular interfaces. This integration enables gene-tailored variant classification grounded in mechanism-specific evidence and supports systematic exploration of genotype–phenotype–function relationships that are not accessible through genome-wide resources alone. By combining these analytical interfaces with educational resources and direct links to patient registries, the GPs function not only as centralized gene-specific knowledge bases but also as integrative platforms that connect clinicians, researchers, families, and patient advocacy organizations.

Accurate variant classification increasingly requires gene-tailored evidence models augmented by expert-guided criteria. Cross-gene analyses consistently show that the weights for a pathogenicity prediction algorithm for variant classification cannot be reliably inferred from genome-wide calibrations alone^24^: even closely related neurodevelopmental genes differ in constraint patterns, functional parameter space, and phenotype–mechanism coupling, such that cross-gene thresholds for these prediction algorithms obscure crucial biological heterogeneity^22,25^. This recognition underlies ClinGen’s move toward gene- and disease-specific ACMG/AMP adaptations, where expert panels define criteria calibrated to known mechanisms, paralog structure, and phenotypic spectrum (ClinGen VCEP Protocol, Version 11, 2023^26^). The GPs operate this principle systematically and at scale: they aggregate and harmonize clinical and molecular datasets that are not available in genome-wide resources to complement population reference datasets, and, when VCEP specifications exist, embed them directly within gene-specific ACMG/AMP workflows. For the *GRIN* and *SCN* GPs, we implemented the published *SCN* and preliminary (to be released soon) *GRIN*-specific refinements^13^ into the automated classifiers. By embedding VCEP-refined criteria within interfaces that organize all gene-relevant evidence, the GPs support interpretation grounded in gene-specific biology rather than generic rule application. They complement expert review by consolidating the information required for multidisciplinary discussions, a format increasingly used in rare-disease genetics to improve VUS adjudication and clinical decision-making^27^.

The GPs provide a research environment in which genotype and phenotype data are harmonized at single-variant resolution, enabling systematic exploration of variant-type- and variant-location-specific clinical patterns and scalable genotype–phenotype modeling. For example, the framework enables the identification of variant-type-specific phenotypic features, including a narrowly constrained age range for seizure onset and offset as well as co-occurrence of schizophrenia in carriers of *GRIN2A* null variants^28,29^. Within the same variant-type–stratified framework, mechanistically informed therapeutic stratification has been suggested, including the selective benefit of L-serine in individuals with GRIN LoF variants and memantine in gain-of-function variants^28,30^. In the *SCN portal*, the harmonized variant–phenotype backbone further enabled the detection of *in silico* and location-dependent patterns that stratify Dravet syndrome and GEFS+ trajectories in individuals with *SCN1A* variants, the two main disease subgroups. This information has already supported the development of early-diagnosis prediction models for *SCN1A-*related epilepsies^31,32^. Extending beyond single genes, gene family-based (“paralog-aware”) mappings across the voltage-gated sodium channel family allowed quantitative incorporation of evidence from related genes. This information revealed gene-family–wide phenotype correlations that are inaccessible to isolated gene-centric analyses^12^.

Beyond genotype–phenotype correlations, the GPs enable systematic interrogation of how variant location and structural context relate to molecular effect and clinical severity. In *GRIN* genes, residue-level mapping demonstrated that spatial distance to agonist and antagonist ligand-binding sites is a strong predictor of both pathogenicity and NMDAR functional impact, providing a basis for NMDAR gene-specific machine-learning models that infer variant pathogenicity and effect direction directly from structural features^23^. Complementary location-based analyses in *SLC6A1* revealed enrichment of loss-of-function variants within specific protein domains and showed that complete loss of GAT1 uptake, encoded by *SLC6A1*, is associated with more severe clinical phenotypes, directly linking residue position, transporter dysfunction, and disease severity^33^. These examples highlight how structural and positional annotation within the GP framework enables mechanistic interpretation of variants and supports quantitative models that connect molecular disruption to patient-level outcomes.

### Limitations

Although the GP framework is designed to be generalizable and has been deployed across diverse gene classes, its depth and interpretability depend on the maturity of gene-specific knowledge and community engagement. GPs developed for genes with limited functional characterization or mechanistic understanding will necessarily provide fewer evidence layers, even though the underlying architecture remains applicable. In addition, certain visualization strategies and evidence metrics reflect assumptions derived from specific molecular contexts and may require validation when extended to other gene classes. While the framework supports the incorporation of emerging variant-relevant dimensions, such as trafficking dynamics or protein–protein interactions, effective expansion requires continued domain expertise and adaptation of the underlying templates. In addition, although the GPs substantially streamline expert review and centralize fragmented data, they are intended as research resources and are not validated for clinical decision-making. Prospective evaluation within diagnostic workflows remains necessary to assess effects on time-to-classification, concordance, reclassification trajectories, and downstream management. Finally, sustained completeness and currency depend on ongoing community contributions, positioning the GPs as collaborative, evolving infrastructures rather than static repositories.

### Conclusion

GPs are a unified, extensible architecture that can transform fragmented clinical, functional, and structural data into interoperable resources for variant classification, and mechanistic analysis in a research context. The framework offers a practical model for developing community-driven, gene-centric knowledge based at scale. As functional assays, population cohorts, and natural history datasets grow, and as computational predictors mature, the GP infrastructure provides a flexible, centralized, and open-access foundation for increasingly automated and comprehensive gene-resolved resources that further strengthen the translation of patient-identified variants into clinically actionable and mechanistically informative insights into rare diseases.

## Methods

### Gene Portal implementation

The Gene Portals (GPs) were implemented in R (v.4.4.0) using the R Shiny framework (https://shiny.posit.com/) (v.1.9.1), enabling R code to be deployed as interactive web applications accessible across all major browsers, including mobile devices. To ensure stable and portable builds, datasets and Shiny application code were packaged into Ubuntu 20.04 LTS Docker^34^ images, deployed on ChromeOS “Lakitu” milestone 113 virtual machines from Google’s stable release channel. All visualizations are generated with ggplot2^35^(v.3.5.2) and plotly^36^(v.4.10.4) for interactive 2D graphics, while protein structures are displayed in 3D using the r3dmol^37^(v.0.1.2) R library. The GPs integrate curated clinical cohorts, functional electrophysiology datasets, and the annotated coding reference set into three user-facing modules (*Clinical Overview*, *Variant Classification*, and *Research*; Fig. 1). These modules provide interactive access to the underlying data resources within a standardized annotation framework. All GPs are publicly available via dedicated domains at https://lalresearchgroup.org.

### Clinical and functional data aggregation and harmonization

Curated clinical and functional data form the foundation of the GPs, informing all three user-facing modules: they provide gene-level clinical summaries in the Clinical Overview module, enable phenotype lookups and support ACMG-based interpretation in the *Variant Classification* module and allow phenotype- and function-based filtering across the *Research* module.

#### Curated patient cohorts

Patient-level data were aggregated from peer-reviewed publications, international registries, clinician-led case series, and institutional databases under formal data-sharing agreements. All datasets were de-identified before integration and were approved by the UTHealth institutional review boards (IRB number: HSC-MS-23-1129). Only cases in which a genetic variant in a target NDD gene and the corresponding clinical phenotypes were documented for the same individual were included. At a minimum, cases included a diagnostic label (*e.g.,* Dravet syndrome). Where available, clinical symptoms such as epilepsy, developmental delay, or movement disorder were annotated, together with quantitative measures such as age at seizure onset. These data elements were captured in standardized formats following established Common Data Elements to ensure comparability across sources. Data were acquired through two models: in some GPs (*GRIN*, *CACNA1A*, *SLC6A1*, *SATB2*), a designated lead collaborator consolidated case-level information prior to transfer; in others (SCN), multiple collaborators contributed datasets directly, and the GP team performed post hoc aggregation. Across sources, potential duplicate cases were systematically identified and excluded. Newly published cases, participants enrolled through disease-specific registries linked to the GPs, and datasets shared by collaborating groups are incorporated on a rolling basis, reflecting the asynchronous availability of curated data from diverse sources. Once retrieved and quality-controlled, contributed datasets are integrated into the GP infrastructure typically within 1-2 days. Public reference resources, including ClinVar and gnomAD, are updated at fixed 6-month intervals. A detailed overview of the clinical cohorts included in each GP is provided in Supplementary Table 2.

#### ClinVar submissions

To complement the curated patient cohorts, which contain detailed harmonized phenotype data, we also incorporated patient-level variants submitted to the public ClinVar database^38^. ClinVar aggregates variant classifications with a phenotype label from clinical laboratories and researchers but typically provides limited or no standardized phenotype information. Variants (accessed May 2024) were downloaded in tabular format from the public FTP site (ftp://ftp.ncbi.nlm.nih.gov/pub/clinvar/) and restricted to those mapped to the genes represented in the GPs. Each variant was harmonized into standardized VCF format and re-annotated using the unified pipeline described below, ensuring direct comparability with cohort-derived data.

#### Functional datasets

Curated functional datasets complement the clinical cohorts and provide experimental insights into the molecular effects of variants. Functional data were aggregated from systematic literature reviews, published studies, and dedicated experimental consortia, and include electrophysiological or molecular readouts from variants tested in heterologous expression systems. Only variants where the raw experimental readouts could be reviewed were included. Reported assays encompassed patch-clamp electrophysiology for sodium and calcium channels, transporter activity assays for GAT1 encoded by *SLC6A1*, and a panel of electrophysiological and expression assays NMDARs encoded by *GRIN1*, *GRIN2A*, *GRIN2B* and *GRIN2D*. Across all GPs, results were curated into harmonized formats and variants classified into functional categories such as gain-of-function, loss-of-function, mixed, or wild-type–like. These functional datasets represent the largest curated resources of their kind for the included genes and are updated on a rolling basis as new experimental results are generated and shared by collaborators. For variants not yet described in the peer-reviewed literature, a panel of experts outside the institution conducting the functional experiments reviewed the raw electrophysiological data to ensure quality. A detailed overview of the functional data included in each GP is provided in Supplementary Table 3.

### Genetic variant standardization and annotation pipeline

To ensure that curated patient cohorts, functionally tested variants, and the full set of all possible missense substitutions can be cross-referenced and jointly analyzed, we developed a unified genetic standardization and annotation pipeline (Fig. 1). Variants not already in VCF format^39^ were first converted using GeneBe^40^. Each variant was then normalized and annotated using our custom ANNOVAR(v.2023 August)^41^ pipeline, which provided HGVS nomenclature at transcript and protein levels, genomic coordinates, exon boundaries, and variant type (*e.g*., missense, frameshift). Coding sequence alignments (MUSCLE^42^, default parameters) were applied to map variants across isoforms and to equivalent positions in paralogous genes from the same gene family, enabling systematic cross-gene comparisons. Paralogs were defined using *HGNC* gene-family assignments^43^ together with downstream subdividing as previously described in Lal et al. 2020^18^. The harmonization step provided a standardized backbone for downstream analyses, including cross-cohort comparisons, paralogous residue mapping, and application of the American College of Medical Genetics and Genomics and the Association for Molecular Pathology (ACMG/AMP) criteria^11^.

#### Genome-wide and gene-specific pathogenicity and functional prediction scores

Each variant was annotated with widely used genome-wide *in silico* pathogenicity predictors, including CADD(v1.4)^17^, REVEL(v1.4)^16^, AlphaMissense (Science 2023 release)^15^, EVE (Nature 2022 release)^44^, MutPred2 (Nat. Comm. 2020 release)^45^, and SpliceAI(v1.4)^46^. These annotations support the application of computational evidence ACMG/AMP criteria PP3 and BP4, providing *in silico* predictions indicative of deleterious or benign effects. While four of the five GPs use REVEL as the default predictor for supporting PP3 and BP4, the *GRIN portal* uses MutPred2 for these criteria in accordance with gene-specific recommendations from the GRIN Variant Curation Expert Panel (VCEP) guidelines^47^. In addition, we annotated a position-level population constraint metric, the missense tolerance ratio^48^, and a paralog conservation score to quantify amino acid conservation within the same gene family^18^. Where available, gene-specific prediction scores were also annotated. We annotated the FuncIon^49^ and SCION^50^ prediction scores, which estimate the functional effect of a variant (gain-versus loss-of-function) on voltage-gated sodium channels in the *SCN portal*. Additionally, we annotated the *GRIN portal* with NMDAR gene-specific pathogenicity and functional prediction scores^23^.

#### Population frequencies and mutational hotspots

Population allele frequencies were obtained from gnomAD(v4.1.0)^6^, along with gene-level intolerance metrics such as missense Z-score and pLI score^51^. To identify protein regions significantly enriched for pathogenic versus control variants, we applied the established approach to calculate pathogenic variant–enriched regions (PERs) using curated patient missense variants as the pathogenic set and all gnomAD missense variants as controls, with parameters set as described in Pérez-Palma et al., 2019^14^. PERs were calculated on the MANE select (Matched Annotation from NCBI and EMBL-EBI)^52^ transcripts for each protein and then mapped to alternative isoforms using the isoform alignments described above. Briefly, missense burden was evaluated in sliding windows of nine amino acids with 50% overlap. For each window, counts of patient and population variants inside versus outside the window were compared using a one-sided Fisher’s exact test (R v4.4.0). Multiple testing correction was performed using a Bonferroni adjustment, accounting for the number of windows across the alignment. Significant windows (adjusted P < 0.05) were merged into contiguous PERs. Identified PERs were used to support the application of the mutational hotspot ACMG/AMP criterion (PM1) across all GPs except the *GRIN portal*. In addition, for the *GRIN portal* we calculated structure-informed 3D missense tolerance ratio (3D-MTR) scores by comparing observed and expected missense versus synonymous variation across spatially neighboring residues defined by protein structural coordinates, thereby quantifying local constraint in three-dimensional space^53^. Intolerant microdomains (iMDs) were defined as contiguous clusters of residues with significantly reduced 3D-MTR values within resolved protein structures and were used to delineate spatial mutational hotspot regions. Identified iMDs were incorporated in the *GRIN portal*, replacing PER-based hotspot definitions for applying the ACMG/AMP PM1 criterion in accordance with gene-specific VCEP guidelines^47^.

#### Variant and domain mapping on the protein sequence and structures

Protein domain annotations were retrieved from the UniProt database^54^ and mapped to the UniProt canonical isoform. Domain positions were cross-referenced with MANE^52^ transcripts to ensure consistent representation across resources. Protein structures were obtained from the Protein Data Bank^55^ (PDB) and the AlphaFold structure server(v2)^56^ for the MANE isoform; for PDB entries, structural coordinates were aligned to the MANE transcript using SIFTS^57^. For visualization within the GP, all variants, including curated patient variants, population variants, and functionally tested variants, were mapped to the MANE Select transcript. This framework enables consistent mapping of variants across protein sequences and three-dimensional structural models.

#### Reference set of all possible coding SNVs

As part of the GP design, we created a comprehensive reference set of all possible single-nucleotide variants (SNVs) in the coding regions of each covered gene, ensuring that any potential variant can be queried and annotated within the GPs. For each gene, all protein-coding transcripts (RefSeq NM, GRCh38; Ensembl BioMart Release 114^58^) were retrieved using the biomaRt (v.2.62.1) R package^59^, with the MANE canonical isoform designated and flagged. From these sequences, we systematically generated all possible codon substitutions, yielding the full set of potential synonymous, missense, and nonsense SNVs. Each variant was processed through the unified annotation pipeline (see above), which added HGVS annotations, population frequencies, constraint metrics, functional domains, structural coordinates, and *in silico* prediction scores. This produced a standardized reference dataset of annotated coding SNVs, enabling consistent variant-level interrogation, comparison across cohorts, and alignment of functional and clinical observations within the broader mutational landscape. Within the VC module, this resource allows users to query any possible SNV and immediately obtain integrated annotations, including semi-automated ACMG/AMP classification (described below). In addition to this SNV reference backbone, the GPs support recognition and interpretation of selected non-SNV variant types at query time. Insertions and deletions within protein-coding regions are parsed to determine coding consequences, including in-frame versus frameshift effects, and exon-level deletions are detected, enabling the automated application of ACMG/AMP criteria such as PVS1. For non-SNV variants queried at the cDNA level, population allele frequencies are retrieved from gnomAD where available, and key attributes, including affected exons and predicted loss-of-function status, are reported. Noncoding variants and splice-site variants outside the canonical ±5 bp window are currently not considered. The framework is designed to support backbone-level annotation of additional non-SNV variant types in future releases.

### Development of an automated ACMG/AMP classification

To operationalize variant classification within the GP, we implemented automated ACMG/AMP criteria assignment with explicit transparency and user control. Criteria were either automatically applied based on available annotations, assigned by user input, or left modifiable. Up to 19 ACMG/AMP criteria (PVS1, PS1, PS2, PS3, PS4, PM1, PM2, PM4, PM5, PM6, PP1, PP2, PP3, PP4, BA1, BS1, BS3, BP4, BP5) are automatically applied based on available data and user input. Quantitative and database-driven criteria were prefilled directly from the harmonized annotations, with details being outlined in Supplementary Table 4. In brief, PM1 from pathogenic variant–enriched regions (PERs) or intolerant 3d microdomains (*GRIN portal*); PM2, BA1, and BS1 from gnomAD allele frequencies; PP3 and BP4 from computational prediction scores; PS3 and BS3 from curated functional data; PS1, PS4 and PM5 from curated patient variants and aggregate variant classifications in ClinVar; PVS1 from the annotated variant type and predicted effect (*e.g*., premature stop, frameshift, canonical splice site); PM4 from variant consequence (*e.g*., in-frame insertion/deletion); and PP2 from missense constraint at the gene or region level (see *GRIN* VCEPs). Criteria requiring user input (*e.g*., PS2/PM6 for *de novo* status, PP4 for phenotype specificity, or segregation evidence) were user-selectable, with embedded guidance provided to support consistent decision-making. For all other non–data-driven or non–user-prompted criteria, users could still manually adjust application and strength levels. Where available, ClinGen VCEP^60^ gene-specific specifications were incorporated to refine evidence thresholds and criteria application. The application of VCEPs is documented in each GP where applicable. Outputs are generated in real time as a five-tier classification (Pathogenic, Likely Pathogenic, VUS, Likely Benign, Benign), with full visibility of applied criteria, rationales, and thresholds.

### Gene Portal-specific customizations

In addition to the core framework implemented across all GPs, each GP was customized to reflect the available data types and community-specific resources. Visualizations in the Clinical Overview and Research sections were adapted to reflect the underlying data structure and the completeness of clinical and functional datasets for each gene. Educational materials were integrated into the GPs in collaboration with the respective family foundations. Short explanatory videos describing the gene or gene family–specific disorders were produced using whiteboard-style animation (VideoScribe v3.9.5, Sparkol 2012; https://www.videoscribe.co/en/download/) and embedded directly within each GP. To improve accessibility and community engagement, each GP also provides direct links to corresponding patient organizations and, where applicable, to active patient registries (*e.g*., the *GRIN portal*).

### User Survey

To evaluate the usability, functionality, and impact of the GPs on variant classification, we conducted a user survey using the REDCap platform^61^. The survey assessed the utility of GPs in interpreting variants in voltage-gated sodium channel genes, NMDAR-encoding genes, *SLC6A1*, and *CACNA1A*, particularly for non-experts. Participation was voluntary, and individuals retained the right to withdraw at any time. Researchers and clinicians with prior genetic knowledge were recruited through in-person outreach at scientific conferences and via email invitations sent to collaborating research centers and department heads. Survey responses were collected between July 6 and August 31, 2024. The survey consisted of five parts: (1) eleven questions assessing demographics and prior experience in variant classification; (2) an initial task to classify two randomly assigned variants (from a pool of four) without using the GP, including documentation of the applied ACMG/AMP criteria and final classification for each variant; (3) viewing a tutorial video introducing the GPs (https://www.youtube.com/watch?v=BObzR8qzeE4); (4) reevaluation of the same two variants using the respective GP, and (5) a follow-up questionnaire comprising 16 questions on usability, accessibility, perceived utility, and suggestions for improvement. The Redcap dictionary of the full survey is available as Supplementary Table 5. The study protocol (IRB: HSC-MS-24-0309) adhered to ethical research standards, including obtaining informed consent, maintaining confidentiality, and providing the right to withdraw.

## Data Availability

Data can be accessed within each GP. The code to reproduce the GPs is available in the corresponding GitHub repositories: https://gitlab.com/neurogenetics/geneportals.

## Declarations

### Ethics approval and consent to participate

Use of de-identified patient-level data integrated from institutional sources was approved by the UTHealth Institutional Review Board (IRB HSC-MS-23-1129). The survey evaluating the impact of the Gene Portals among medical professionals was approved under a separate UTHealth IRB protocol (HSC-MS-24-0309). Published and registry-derived data were used in accordance with the originating source terms and applicable governance.

## Acknowledgement

We gratefully acknowledge Felicia Mermer, Maina Kava, Thomas Balslev, Marc Engelen, Marwan Shinawi, Katherine A. Bosanko, Anne Ducros, Kristin Baranano, Elisabetta Indelicato, Julia Koh, Sooyeon Jo, Anna Abuli Vidal and Line Futtrup for their valuable contributions to the development, refinement and community integration of the GPs. We thank the individuals and families who participated in research studies and generously provided feedback that informed us about the design and functionality of the GPs. We further acknowledge the support of all patient advocacy organizations (Supplementary Table 1) and community partners whose sustained collaboration enabled cross-GP dissemination and implementation of this framework.

## Availability of data and materials

Data can be accessed within each GP; downloadable materials are described in the repository, while patient-level data are viewable in the GPs and available upon request of the corresponding author. The code to reproduce the GPs is available in the GitHub repository: (https://gitlab.com/neurogenetics/geneportals).

## Conflict of interests

A. Brunklaus has received honoraria for presenting at educational events, advisory boards and consultancy work for Biocodex, Encoded Therapeutics, Jazz Pharma, Servier, Stoke Therapeutics, and UCB. S. Boesch has served as a consultant for VICO Therapeutics, Reata Pharmaceuticals, and Biogen; has participated on advisory boards for Biogen, Reata Pharmaceuticals, and Biohaven; and has received honoraria from Ipsen, Merz, Reata Pharmaceuticals, and Biogen. AW, MT, and SF are current or past employees of BioMarin Pharmaceutical Inc K. Johannesen is on the advisory board of SLC6A1 Connect Europe. SY.B. is a member of the European Reference Network for Rare Neurological Diseases (Project ID No. 739510).

## Funding

Funding for this work was provided by the German Federal Ministry for Education and Research (BMBF, Treat-ION, 01GM1907D) to D.L., T.B., and P.M., by the BMBF (Treat-Ion2, 01GM2210B, 01GM2210A) to P.M and H.L., by the Fonds Nationale de la Recherche in Luxembourg (FNR, Research Unit FOR-2715, INTER/DFG/21/16394868 MechEPI2) to P.M., by the Chilean National Agency for Research and Development to E.P.P., (ANID) Fondecyt grant 1221464 to E.P.P., by the Dravet Syndrome Foundation (grant number, 272016) to D.L, the by NIH NINDS (Channelopathy-Associated Epilepsy Research Center, U54-NS108874) to A.L.G., J.Q.P., C.G.V., I.H., and D.L., the Agence Nationale de la Recherche - France (Initiative of Excellence Université Côte d’Azur ANR-15-IDEX-01) to SC and MM, 23% the MRC (MR/T002506/1) to M.F., the CureGRIN Foundation to M.F.by the NIH-NINDS (NS111619 SFT), the NIH-NIMH (MH127404 H.Y), NICHD (HD082373 H.Y), the GRIN2B Foundation (H.Y), GRIN Therapeutics (S.F.T and S.J.M), Austin’s Purpose (S.F.T), SFARI (732132 to S.F.T), by the University Research Committee (Emory URC to H.Y), by Imagine, Innovate and Impact (I^3^) Awards from the Emory University School of Medicine and through the Georgia CTSA NIH award (UL1-TR002378; H.Y), by the National Institute of Health grants S10MH133644 (J.Q.P.), NS108874 (J.Q.P.), MH131719 (J.Q.P.), MH129722- 02 (M. D.), and Stanley Center for Psychiatric Research (J.Q.P. and M.D.), and by grants from CACNA1A foundation (J.Q.P.) and the Ladders to Cures Scientific Accelerator of the Broad Institute of MIT and Harvard (J.Q.P.). The *CACNA1A portal* development was funded in part by the Chan Zuckerberg Initiative Rare as One grant. A.P. received support for Simons Searchlight by the Simons Foundation.

## Author’s Contributions

Conceptualization: T.B., D.L., C.K., Ma.Mac., E.P.P., A.P., M.J.D., Y.A.Z., J.R.L.; Data collection and curation: I.K., Suy.K., S.J.M., K.M.J., S.S., H.Y., R.E.P., Suk.K., I.H., J.Q.P., M.F., L.W., D.J.A.W., E.K., D.B., S.Z., C.M.B., Ma.Man., Sa.C., A.I., K.G., S.D.W., K.V.E., K.H., J.Q.K., C.G., A.W., M.T., S.F., A.T.D., A.A.S., Art.B., A.Sh., Arj.B., MJ.H., D.K.P., L.D., D.L., T.R.B., E.E.O.H., A.H., K.O., T.L.H., P.S., T.A.B., V.S., K.P., A.R., W.C., A.P., L.L.S., S.M.R., St.C., L.S., Sy.B., O.W., A.J.P., E.C., N.B., M.P.H., A.K., C.G.V., G.Q.Z., And.B., S.F.T., R.S.M., J.R.L. Front and backend development: T.B., C.K., Ma.Mac., Suy.K., A.St., G.T., R.S.D; GP design (feedback): T.B., D.L., C.K., Ma.Mac., S.J.M., I.K., R.S.D., I.H., P.M., M.A.M., V.H., L.S.M., A.F., L.M., S.M., P.F., J.N., M.N., Y.A.Z., And.B., S.F.T., R.S.M., I.K.; Writing-original draft: T.B., D.L.; Writing-editing: A.L.G., I.K., Suy.K., G.T., E.P.P., R.S.D., H.Y., R.E.P., Suk.K., Su.B., I.H., J.Q.P., M.F., L.W., D.J.A.W., E.K., C.M.B., H.L., P.M., A.I., L.M., A.Sh., MJ.H., T.R.B., T.L.H., T.A.B., W.C., A.P., Sy.B., A.K., C.G.V., M.N., And.B., S.F.T., R.S.M., J.R.L., D.L., T.B.

## Notes

### Author Declarations

Ethical approval for this study was granted by the Institutional Review Board of The University of Texas Health Science Center at Houston (UTHealth Houston) under two protocols: HSC-MS-23-1129 ('Delineating the clinical spectrum of rare epilepsies'), covering clinical and research data aggregation, and HSC-MS-24-0309 ('Evaluation of the Gene Portals Impact on Genetic Variant Interpretation Among Medical Professionals'), covering the survey evaluating the Gene Portals platform among medical professionals.

