## Supplementary Data for "Gene Portals: A Framework for Integrating Clinical, Functional, and Structural Evidence into Rare Disease Variant Classification"

### Supplements

#### Supplementary Note 1 | User Feedback and Global Reach

To assess the real-world impact of GPs, we conducted a user survey among clinicians and researchers (see Methods for details). Among 62 respondents who were not contributors to the portals (50 clinicians and 12 researchers) 68% reported that they were “likely” or “very likely” to use the GPs to support variant classification in their workflow (Supplementary Figure 1A), and 62% indicated that the GPs accelerate variant classification (Supplementary Figure 1B). A similar proportion (62%) of respondents stated that the portals provided novel information that improved their classification decisions (Supplementary Figure 1C), suggesting that the integrated, gene-specific evidence layers provide information beyond standard resources used for variant interpretation. Web analytics further supported their utility. From May to October 2025, the portals collectively attracted an average of 650 monthly active users, with the highest engagement observed for the GRIN portal, followed by the SCN, SLC6A1, CACNA1A and SATB2 portals (Supplementary Figure 1D). Access data shows global adoption of the GPs suggesting wide applicability (Supplementary Figure 1E). Together, these observations indicate that the GPs are not only technically capable of integrating multimodal evidence but are also being adopted by the intended user communities as practical tools for variant classification, mechanistic interpretation, and collaborative research.

### Variants used in the classification survey

Within the survey participants received 2 out of four randomly selected variants to classify first without and later with the use of the GP. The variants were selected to represent different genes, inheritance patterns and clinical presentations commonly encountered during variant classification. The four variants with their clinical description are listed below:

1) NM_001165963 (SCN1A):c.2362G>A (p.Glu788Lys) - Heterozygous - Inheritance unknown
   Description: A young female who developed seizures at age 15 months in the setting of illness. Currently experiences absence and generalized tonic-clonic seizures that are triggered by heat or strenuous activity. History is also positive for developmental delay. The Inheritance of the SCN1A variant is unknown.

2) NM_001040142 (SCN2A):c.5146T>C (p.Trp1716Arg) - Heterozygous - *de novo*
   Description: An older teenager with autism spectrum disorder, global developmental delay (non-verbal), and self-injurious behavior. History of 1 seizure at age 11 and EEG that showed multifocal epileptiform discharges.

3) NM_003042 (SLC6A1):c.131G>A (p.Arg44Gln) - Heterozygous - Inheritance Unknown
   Description: A female child with intellectual disability, speech delay, behavioral issues, and absence seizures that began at 1 year of age. She is taking valproic acid and Depakote, but continues to have daily seizures.

4) NM_000834 (GRIN2B):c.1306T>C(p.Cys436Arg) - Heterozygous - *de novo*
   Description: A male young adult with autism spectrum disorder, developmental delay, ADHD, OCD, and absence seizures. He has a history Pediatric Autoimmune Neuropsychiatric Disorders Associated with Streptococcal Infections (PANDAS) at age 13, which led to worsening of behavior issues.

### Supplementary Figures

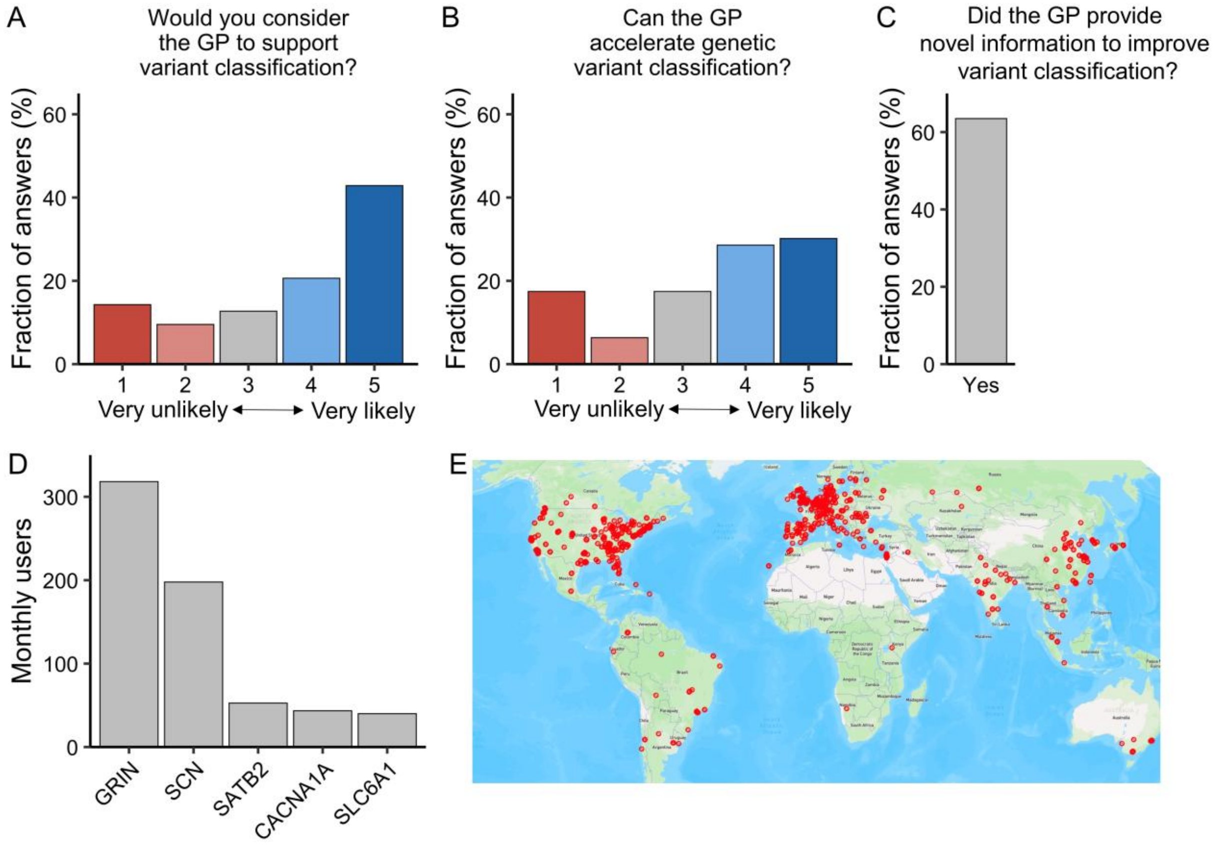


**Supplementary Figure 1 | Global usage and user-reported impact of the GPs. (A)** Likelihood of using the GPs for variant classification, shown on a 5-point Likert scale. Most users rated them as "likely" or "very likely." **(B)** User ratings of whether the portals accelerate variant classification, with responses skewed toward higher values. **(C)** Proportion of respondents reporting that the portals provided novel information that improved classification, with the vast majority answering "yes." **(D)** Monthly user numbers for each portal showing highest engagement with *GRIN*, followed by *SCN*, *SLC6A1*, *CACNA1A*, and *SATB2*. **(E)** Geographic distribution of users worldwide, with the highest concentrations in North America, Europe, and Asia.

### GRIN-Portal: Variant Analysis Report

**Date:** 2026-03-02 15:41

**No medical advice.** The contents have been carefully checked and created to the best of our knowledge. But for the information presented and conclusions made here is no claim to completeness, timeliness, quality, and accuracy. We cannot accept any responsibility for any damage caused by reliance on or use of the contents of this report.

#### Variant Summary

**Gene:** GRIN2B
**Transcript:** NM_000834.5

**Variant input:** p.Gly820Ala

**Inheritance Pattern:**
**Zygosity:** Heterozygous

**Variant classification (optionally user customized)[1]:** Pathogenic

**Applied Criteria:** PM1(Moderate), PM2(Supporting), PM5(Moderate), PS4(Moderate), PP2(Supporting)

**Functional Consequence[2]:** Likely LoF

[1] ACMG Criteria were automatically annotated according to standard ACMG Variant Interpretation Guidelines (https://pubmed.ncbi.nlm.nih.gov/25741868/) (*GRIN1*, *GRIN2A*, *GRIN2D*) and the preliminary ClinGen GRIN Expert Panel Specifications (https://clinicalgenome.org/affiliation/50078/) to the ACMG/AMP Variant Interpretation Guidelines (to be released soon). Classification not to be used in a medical context.
[2] **Likely GoF/LoF**: One or more changes with high confidence and no conflicts in direction of change. **Possible LoF/GoF**: Either two or more changes with moderate confidence and no conflicts in direction of change or one change of moderate confidence and >2.5-fold or <0.4-fold change in synaptic or non-synaptic charge transfer or conflicting changes of moderate/high confidence and >2.5-fold or <0.4-fold change in synaptic or non-synaptic charge transfer. **No detectable effect**: No detectable functional changes for any parameters or only one change with moderate confidence and a change in synaptic and non-synaptic charge transfer between 0.4 and 2.5-fold. **Indeterminant**: Conflicting changes in opposite functional direction and a change in synaptic and non-synaptic charge transfer between 0.4 and 2.5 fold. **Indeterminant***: Variants in which all six assays were completed but the current amplitude of NMDAR responses in transfected HEK cells recorded under voltage clamp were too small to accurately determine the deactivation tau, and thus we cannot determine the overall outcome (Xu et al., CMLS, 2024).* **Likely LoF**: Large decrease in response amplitude that precludes other parameter assessments plus evidence for protein synthesis.

#### Disease Summary

*GRIN2B*-related disorder is characterized by mild-to-profound developmental delay / intellectual disability in all affected individuals. Other common manifestations are epilepsy, muscular hypotonia, movement disorders, spasticity, feeding difficulties, and behavior problems. A subset of individuals shows a malformation of cortical development consisting of extensive and diffuse bilateral polymicrogyria. More details can be found on GeneReviews. (https://www.ncbi.nlm.nih.gov/books/NBK501979/)

#### Detailed analysis

##### Population data

**Applied criteria:** PM2 (Supporting)  **Criteria not applied:** BA1, BS1, BS2

gnomAD:

Alelle frequency: 0
Alelle count: 0

gnomAD highest subpopulation allele frequency:

0 (No subpopulation defined)

##### Patient data

**Applied criteria:** PS1 (Strong), PM5 (Moderate)  **Criteria not applied:** PP5

Individuals with the same variant have been observed 8 times in our patient cohort.
Individuals with the same amino acid exchange, but different nucelotide exchange have been observed 0 times in our patient cohort.
Individuals with a different amino acid exchange have been observed 4 times in our patient cohort.

The variant was reported in ClinVar as Pathogenic (https://www.ncbi.nlm.nih.gov/clinvar/variation/208643).
A variant with a different amino acid exchange was reported in ClinVar as Pathogenic (https://www.ncbi.nlm.nih.gov/clinvar/variation/234635).

#### Computational predictions

**Applied criteria:** None **Criteria not applied:** PP3, BP4

MutPred2:NA

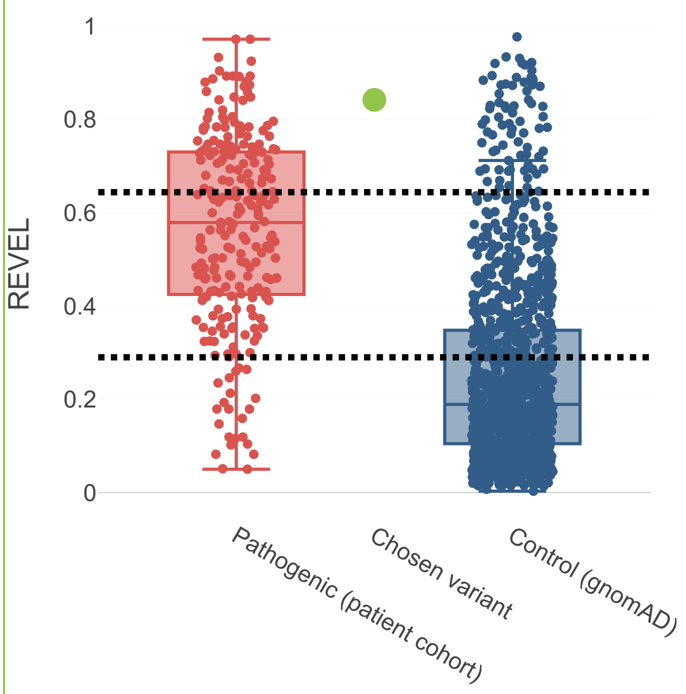


REVEL:0.842

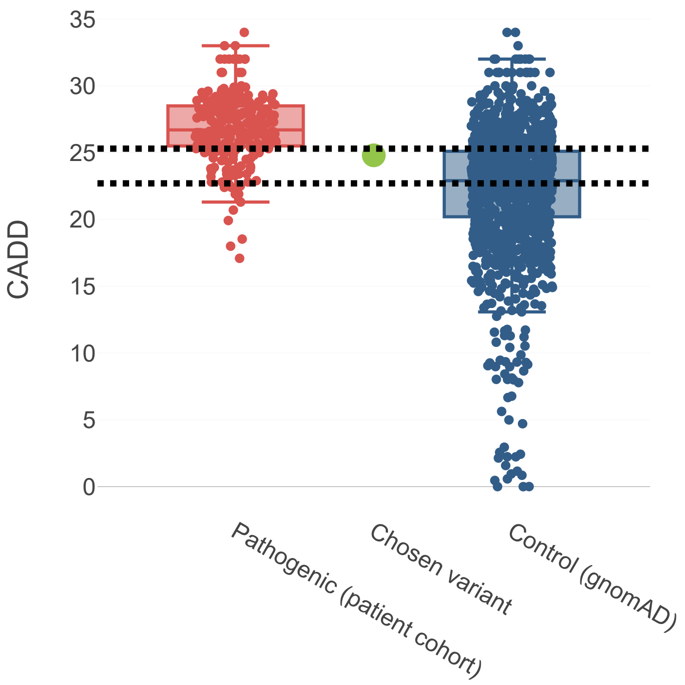


CADD:24.8

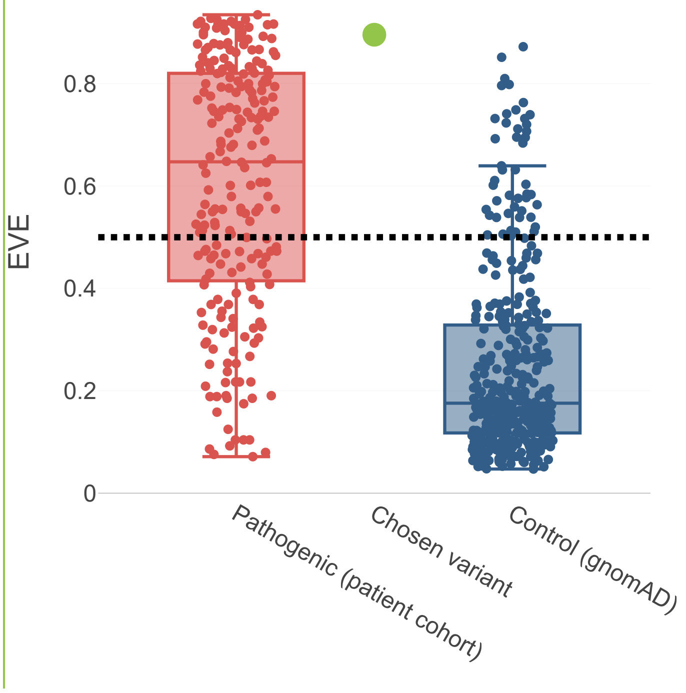


EVE:0.896

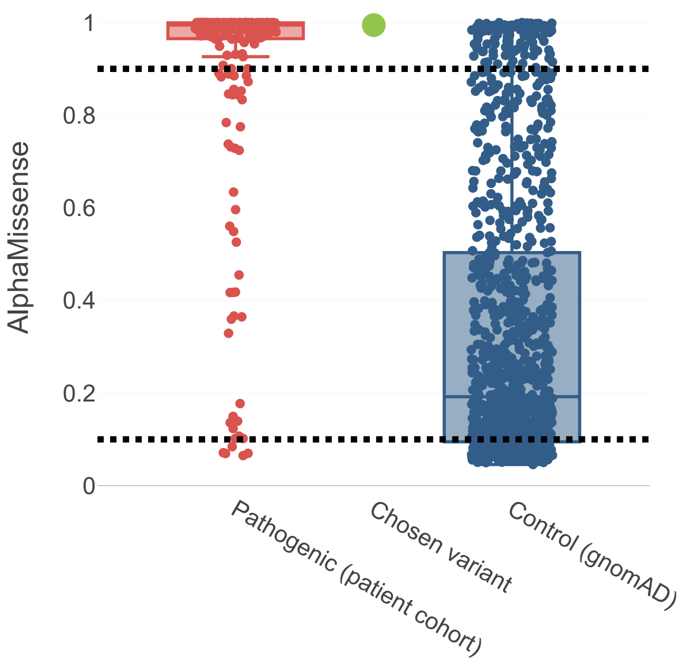

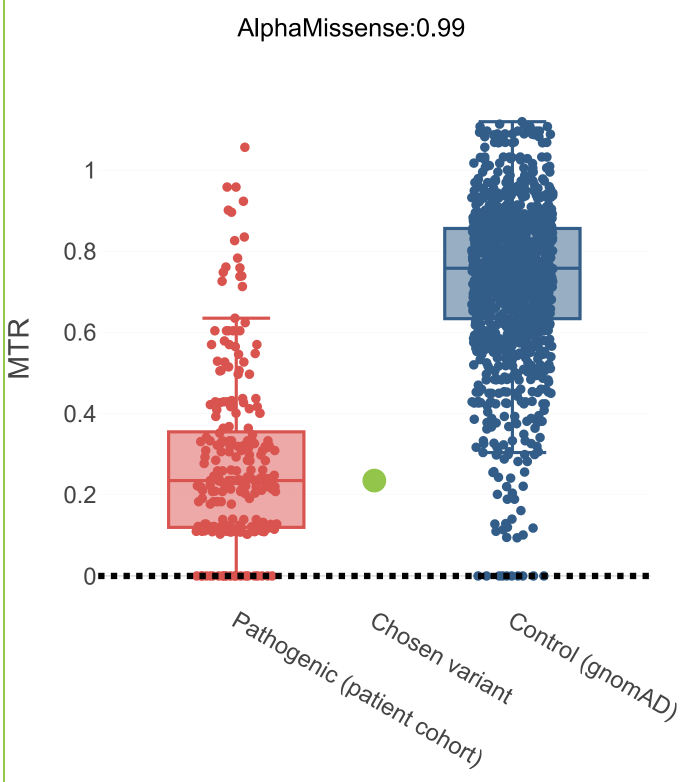


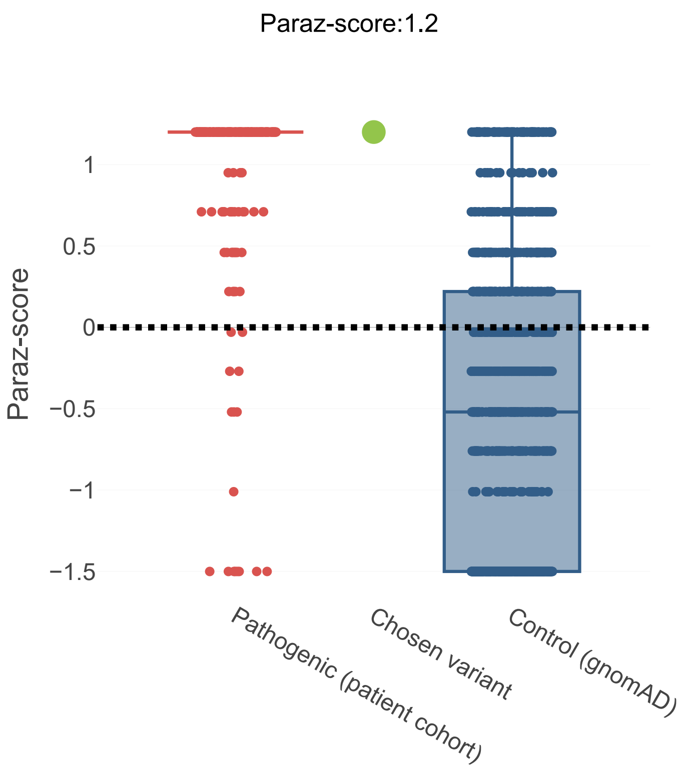


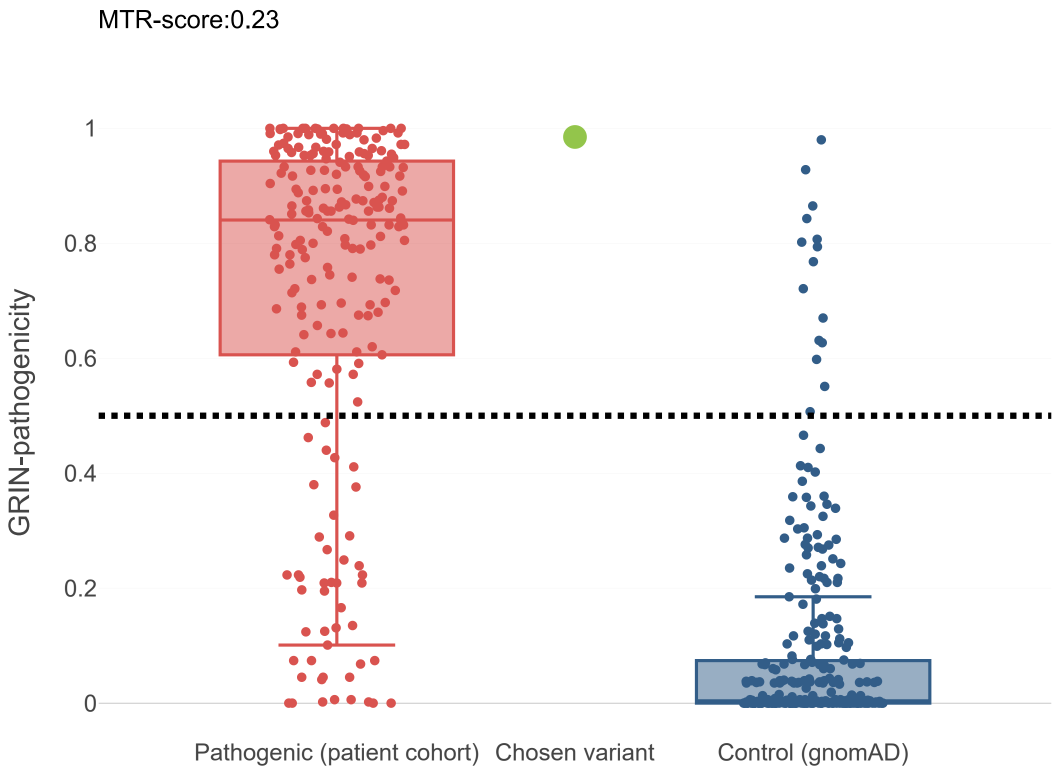


#### Hotspot data

**Applied criteria:** PM1 (Moderate) **Criteria not applied:** None

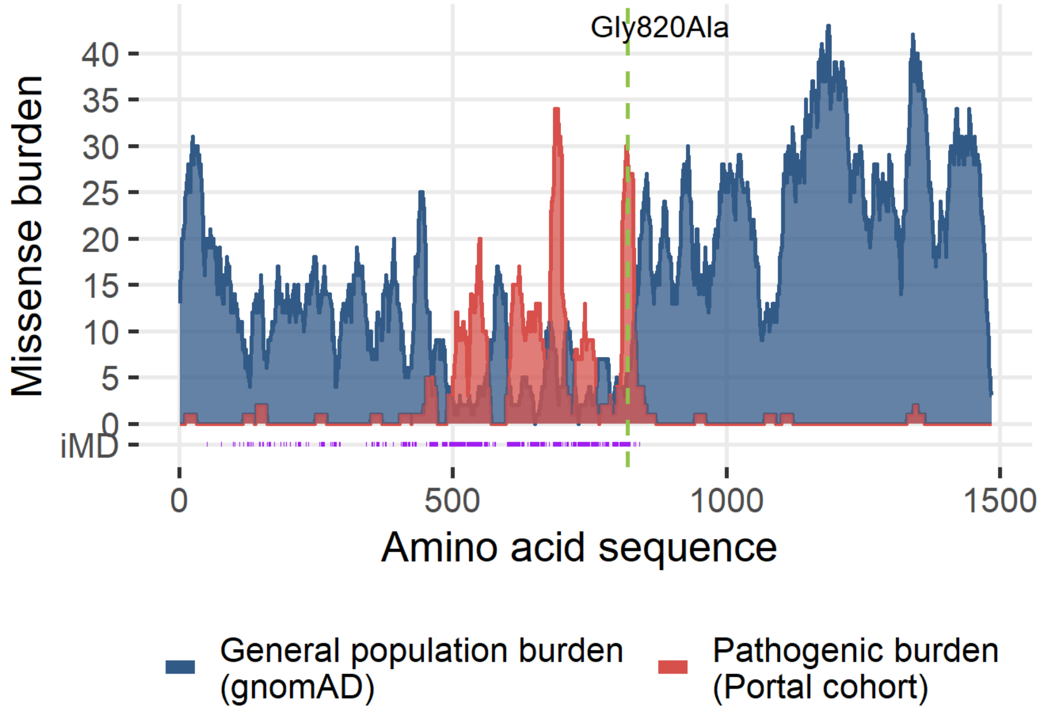


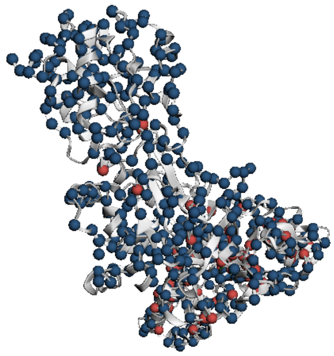

Functional data (Detailed net changes of the NMDA receptor activity - in vitro assay)

The fold changes in NMDA receptor function were assessed in vitro and compared to wildtype receptors. There are no variant fold changes observed leading towards an increase in NMDA receptor activity. The variant fold changes observed leading towards a decrease in NMDA receptor activity are: Magensium:0.9; PH:0.8; Open probability:0.5; Surface expression:0.8; TauW:0.3. The calculated fold synaptic charge transfer by the variant compared to wildtype controls was 0.14.The calculated fold non-synaptic charge transfer by the variant compared to wildtype controls was 0.46.

#### Methods

General Aspects

The GRIN Portal is a coalition of investigators seeking to aggregate and harmonize data generated to study GRIN-related disorders, and to make summary data interactively accessible for the wider scientific community, while providing educational resources for everyone. This automatically generated report provides an overview on the evaluation of a selected GRIN variant in the context of this website. We combine multiple expert-curated resources on genetic, clinical, and molecular data.

##### Limitations

The analysis has been performed in a research setting. The contents of this report have been carefully checked and created to the best of our knowledge. But for the information presented here is no claim to completeness, timeliness, quality and accuracy. No responsibility can be accepted for any damage caused by reliance on or use of the contents of this report. We object to any commercial use and disclosure of data.

In case there are any further questions, please do not hesitate to contact us: (mailto:).
